# Impaired incretin homeostasis in non-diabetic moderate-severe CKD

**DOI:** 10.1101/2023.12.15.23300050

**Authors:** Armin Ahmadi, Jorge Gamboa, Jennifer E. Norman, Byambaa Enkhmaa, Madelynn Tucker, Brian J. Bennett, Leila R. Zelnick, Sili Fan, Lars F. Berglund, Talat Alp Ikizler, Ian H. de Boer, Bethany P. Cummings, Baback Roshanravan

## Abstract

**Background:** Incretins are regulators of insulin secretion and glucose homeostasis that are metabolized by dipeptidyl peptidase-4 (DPP-4). Moderate-severe CKD may modify incretin release, metabolism, or response.

**Methods:** We performed 2-hour oral glucose tolerance testing (OGTT) in 59 people with non-diabetic CKD (eGFR<60 ml/min per 1.73 m^2^) and 39 matched controls. We measured total (tAUC) and incremental (iAUC) area under the curve of plasma total glucagon-like peptide-1 (GLP-1) and total glucose-dependent insulinotropic polypeptide (GIP). Fasting DPP-4 levels and activity were measured. Linear regression was used to adjust for demographic, body composition, and lifestyle factors.

**Results:** Mean eGFR was 38 ±13 and 89 ±17ml/min per 1.73 m^2^ in CKD and controls. GLP-1 iAUC and GIP iAUC were higher in CKD than controls with a mean of 1531 ±1452 versus 1364 ±1484 pMxmin, and 62370 ±33453 versus 42365 ±25061 pgxmin/ml, respectively. After adjustment, CKD was associated with 15271 pMxmin/ml greater GIP iAUC (95% CI 387, 30154) compared to controls. Adjustment for covariates attenuated associations of CKD with higher GLP-1 iAUC (adjusted difference, 122, 95% CI -619, 864). Plasma glucagon levels were higher at 30 minutes (mean difference, 1.6, 95% CI 0.3, 2.8 mg/dl) and 120 minutes (mean difference, 0.84, 95% CI 0.2, 1.5 mg/dl) in CKD compared to controls. There were no differences in insulin levels or plasma DPP-4 activity or levels between groups.

**Conclusion:** Incretin response to oral glucose is preserved or augmented in moderate-severe CKD, without apparent differences in circulating DPP-4 concentration or activity. However, neither insulin secretion nor glucagon suppression are enhanced.

**Significance statement:** CKD is associated with metabolic and physiological disturbances including disturbed glucose and insulin homeostasis. Incretin hormones are potent regulators of insulin secretion and glucose metabolism; however, determinants and potential consequence of incretin response in CKD are incompletely understood. This study revealed that total incretin levels and incretin response during oral glucose tolerance testing (OGTT) were significantly higher among patients with moderate-severe non-diabetic CKD compared to healthy. Unlike in healthy individuals, increased incretin response was not correlated with insulin and C-peptide response to OGTT in CKD. These differences coincided with persistently greater glucagon levels in response to physiological stimuli in CKD. Disruption in the incretin system and glucagon dynamics may contribute to metabolic disturbances in moderate-severe CKD.

**Visual abstract:** 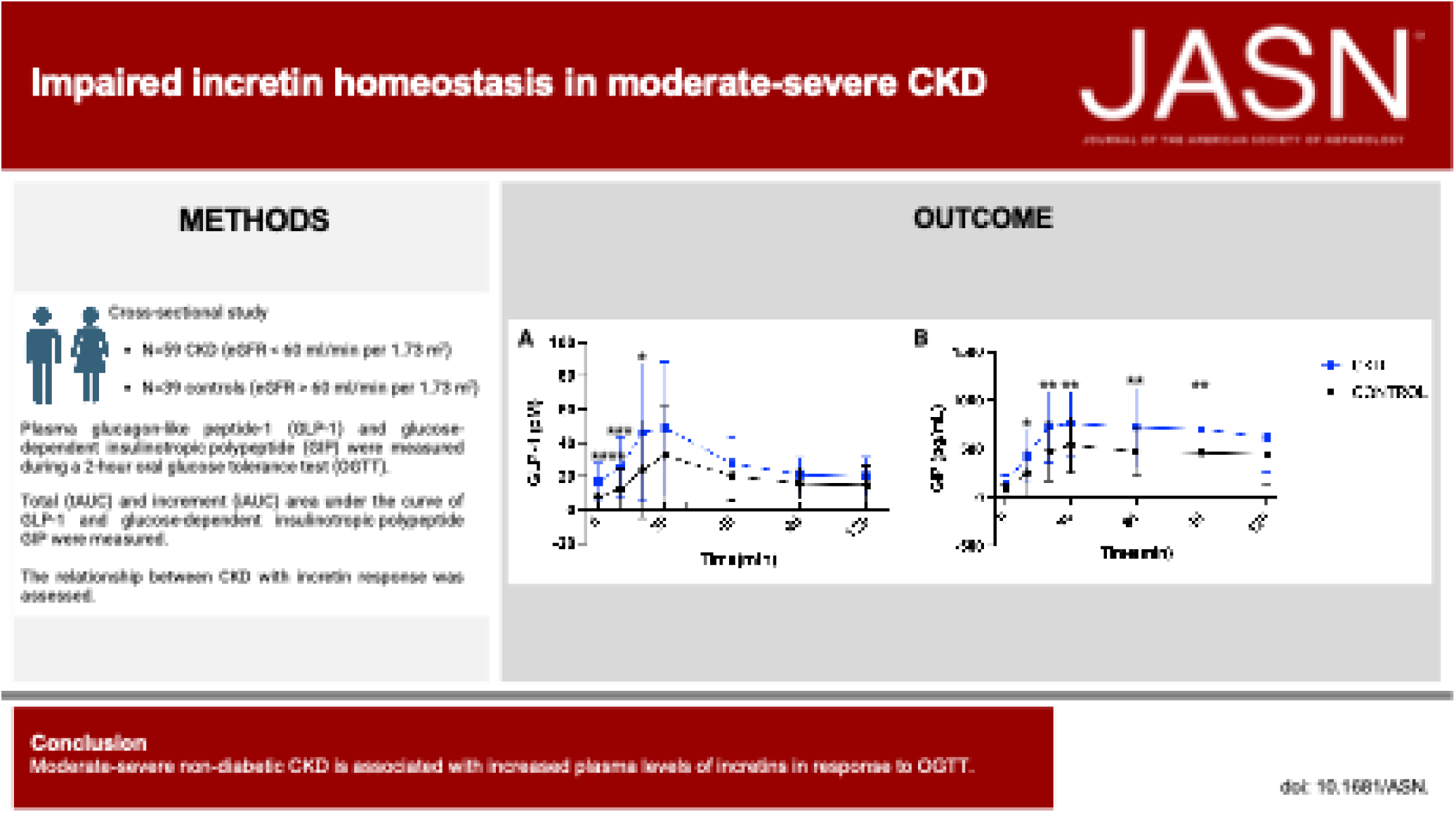

## Introduction

Non-diabetic chronic kidney disease (CKD) is associated with metabolic dysregulation including disrupted insulin and glucose homeostasis ^1–3^. Factors contributing to CKD-associated glucometabolic complications include increased inflammation^4^ and hyperglucagonemia^5^. Prior studies in CKD using hyperinsulinemic-euglycemic clamp and oral glucose tolerance testing have demonstrated lower insulin clearance and insulin sensitivity that is not compensated for by enhanced insulin secretion, leading to a high prevalence of glucose intolerance ^3^. An impaired response of incretin, a key regulator of insulin secretion and glucose homeostasis, could be an important mechanism contributing to inadequate insulin secretion in CKD. However, understanding of how CKD impacts postprandial incretin secretion is limited. This knowledge is key to understanding any potential heterogeneity in response to incretin analogues in the CKD population.

Incretin hormones are secreted by the gut in response to nutrient intake and promote glucose-stimulated insulin secretion ^6^. The two main incretin hormones are glucagon-like peptide-1(GLP-1) and glucose-dependent insulinotropic polypeptide (GIP) secreted by the enteroendocrine L and K cells, respectively ^7,8^. Together, GLP-1 and GIP account for up to 70% of postprandial insulin secretion (incretin effect) in healthy individuals ^9^. While patients with type 2 diabetes are thought to have an impaired incretin effect ^10^, little is known about the independent effect of CKD on the response of the incretin peptides to nutrient ingestion and the islet endocrine cells’ response to them. Both incretins similarly mediate the gastrointestinal glucose-dependent stimulation of insulin secretion. However, the incretins have opposing effects on glucagon secretion with GLP-1 suppressing^11^, and GIP stimulating glucagon secretion ^12^. Whether and how GLP-1 and GIP in combination impact postprandial glucagon suppression in CKD remains unknown. Additionally, understanding the impact of CKD on dipeptidyl peptidase-4 (DPP-4), a ubiquitous enzyme that inactivates incretin hormones, to impact glucagon and insulin release and thus glucose homeostasis is lacking ^13^.

The current study investigates postprandial incretin hormone levels and their determinants using a standardized oral glucose tolerance test (OGTT) comparing non-diabetic patients with CKD and controls. We first describe the association of the presence and severity of kidney disease with circulating concentrations of incretin hormones in both fasted and postprandial states. We separately investigate the association of postprandial circulating incretin hormones with insulin, c-peptide, and glucagon levels during an OGTT by CKD status. We hypothesized that non-diabetic CKD is associated with reduced incretin hormone release and impaired glucagon suppression that contribute to glucometabolic complications underlying heightened cardiometabolic risk in CKD.

## Methods

### Study population and study design

The Study of Glucose and Insulin in Renal Disease (SUGAR) was a cross-sectional study of moderate-severe non-diabetic CKD. A total of 98 participants were recruited for this study among which 59 had CKD (eGFR < 60 ml/min per 1.73 m^2^) and 39 were controls (eGFR > 60 ml/min per 1.73 m^2^), frequency matched on age, sex, and race. Exclusion criteria for both groups included age <18 years, a clinical diagnosis of diabetes, maintenance dialysis or fistula in place, history of kidney transplantation, use of medications known to reduce insulin sensitivity (including corticosteroids and immunosuppressants), fasting serum glucose ≥126 mg/dl, and hemoglobin <10 g/dl. All enrolled participants attended a screening visit, at which eligibility was assessed and written informed consent was obtained. Serum biomarkers of kidney function were measured in fasting blood. A more detailed description of the study design, recruitment, and enrollment has been published previously ^3,14^.

### CKD classification

Serum creatinine and cystatin C (Gentian) were measured in fasting serum collected immediately prior to the hyperinsulinemic-euglycemic clamp using a Beckman DxC automated chemistry analyzer. Primary analyses used GFR estimated using the CKD-EPI Creatinine-Cystatin C Equation (2012) ^15^ to follow precedent of the original eligibility criteria, categorizations, and analyses. Sensitivity analyses were performed using the more recent race-neutral CKD-EPI Creatinine-Cystatin C Equation (2021) ^16^.

### Oral glucose tolerance test and hyperinsulinemic-euglycemic insulin clamp

A standard 75g OGTT was performed approximately one week after the hyperinsulinemic-euglycemic insulin clamp. Plasma glucose, insulin, total GLP-1, and total GIP concentrations were measured at –10, -5, 0, 30, 60, 90, and 120 minutes. We averaged -10 to 0 time points to generate baseline fasting values. Plasma glucagon levels were measured at 0, 30, and 120 minutes. The postprandial incretin hormone responses were calculated as the area under the curves(AUC) using the trapezoid rule and evaluated both as total AUC (tAUC) and incremental AUC (iAUC). Glucose iAUC and 2-hour plasma glucose were calculated as a measure of glucose tolerance. Insulinogenic index was used to quantify the difference in plasma insulin divided by the difference in plasma glucose from baseline to 30 minutes of the OGTT. Clamp insulin sensitivity and Matsuda index were the primary and secondary measures of insulin sensitivity. Details of the clamp and OGTT procedures have been published previously ^17,18^.

### Measurement of GLP-1, GIP, glucagon, insulin, glucose, C-peptide, DPP-4, and inflammatory biomarkers

Plasma samples were assayed for total GLP-1 and total GIP using multiplex electrochemiluminescence (Meso Scale Discovery, Rockville, MD, USA). Average intra-run concentration coefficients of variation for GIP and GLP-1 were 8.3% and 2.7% (high control), 4% and 2.5% (medium control), and 11% and 3.6% (low control) respectively. Plasma glucagon was measured by ELISA (Mercodia). DPP-4 antigen concentration was determined by ELISA (eBioscience). Average intra-run concentration coefficients of variation for glucagon were 2.1% (high control), 14% (medium control), and 6.3 %(low control). Blood glucose concentrations were measured using the glucose hexokinase method (Roche Module P Chemistry autoanalyzer; Roche, Basel, Switzerland) and blood insulin concentrations were measured using 2-site immune-enzymometric assay (Tosoh 2000 Autoanalyzer). C-peptide concentrations were determined using a standard double-antibody radioimmunoassay (Diagnostic Products Corporation, Los Angeles, CA, USA). DPP-4 activity was assayed by incubating plasma with a colorimetric substrate, l-glycyl-l-prolyl p-nitroanilide, hydrochloride (Sigma), at 37°C. Serum inflammation biomarkers were measured in the fasting blood. CRP was measured with a Beckman Coulter (USA) DxC chemistry analyzer. Serum TNF-α, IL-6, IFN-γ, and IL-1*β* were performed using commercial multiplex electroluminescence assays (Meso Scale Discovery, Rockville, MD, USA). All assays were performed in duplicate.

### Covariates

Demographic and medical history of participants were self-reported. Cardiovascular disease (CVD) was defined as a physician diagnosis of myocardial infarction, stroke, resuscitated cardiac arrest, or heart failure or a history of coronary or cerebral revascularization. The Human Activity Profile (HAP) maximum activity score was used to quantify physical activity. Food intake was recorded using three days of prospective food diaries analyzed with Nutrition Data System for Research software. Body composition was measured by DXA (GE Lunar or Prodigy and iDXA; EnCore Software versions 12.3 and 14.1; GE Healthcare, Waukesha, WI).

### Statistical analysis

To compare plasma incretin levels by CKD status during OGTT, we used linear regression adjusted for potential confounders including age, sex, smoking status, fat-free mass, fat mass, calorie intake, physical activity, and CVD. All clinical data was checked for normality. Spearman correlation coefficient was used to evaluate univariable relationship between kidney function and incretin levels during the OGTT. Total and incremental AUCs were used to evaluate total incretin hormone levels and incretin hormone responses during the OGTT, respectively. The rate of acute incretin peripheral response was calculated using the difference of plasma incretin levels at baseline and 30 minutes post OGTT and over time. Linear regression adjusted for confounders was used to investigate the association of CKD status with incretin levels and incretins with measures of insulin resistance, plasma insulin concentrations, and plasma inflammatory biomarkers. Analyses were conducted using R version 4.2.2 ^19^. Boxplots and scatterplots were made using GraphPad Prism version 10.0.0 (GraphPad Software, Inc., San Diego, California).

### Study approval

The procedures in the study and informed consent forms were reviewed and approved by the University of Washington Human Subjects Division (HSD). All participants provided written informed consent.

## Results

### Characteristics of the study participants

The study included a total of 98 total participants, of whom 59 had CKD (eGFR <60 ml/min per 1.73 m^2^) and 39 were healthy controls (eGFR ≥60 ml/min per 1.73 m^2^). The mean (± SD) age among CKD participants was 63.6 ± 13.9 years, 51% were female, and 22% self-identified as black. Mean (range) eGFR was 37.6 (9.5 to 59.5 ml/min per 1.73 m^2^) compared to 88.8 (61 to 117 ml/min per 1.73 m^2^) among controls (**Table 1**). Compared with controls, participants with CKD were more likely to have cardiovascular disease, to be smokers, be less physically active, have higher body weight, fat mass, and plasma inflammatory markers, and have lower daily calorie intake (**Table 1**).

**Table 1.** Characteristics of participants in the Study of Glucose and Insulin in Renal Disease.

| Characteristics | Controls | CKD |
| --- | --- | --- |
| **Number** | 39 | 59 |
| **Demographics** |  |  |
| Age | 61.0 (12.4) | 63.6 (13.9) |
| (%) Female | 17 (44) | 30 (51) |
| (%) Race |  |  |
| White | 34 (87) | 41 (69) |
| Black | 4 (10) | 13 (22) |
| Asian/Pacific Islander | 1 (3) | 5 (8) |
| **Medical history and lifestyle** |  |  |
| History of CVD | 2 (5) | 19 (32) |
| Currently smoking | 3 (8) | 10 (17) |
| Physical activity, HAP score | 83.5 (8.7) | 76.8 (9.5) |
| Calorie intake, kcal | 2047.9 (556.4) | 1758.2 (540.8) |
| **Physical characteristics** |  |  |
| BMI (kg/m<sup>2</sup>) | 27.5 (6.3) | 30.2 (6.0) |
| Body weight (kg) | 82.1 (20.6) | 88.1 (19.8) |
| Fat-free mass (kg) | 56.1 (13.1) | 53.7 (11.7) |
| Fat mass (kg) | 27.1 (14.0) | 31.9 (11.6) |
| **Laboratory data** |  |  |
| Serum creatinine (mg/dl), median (IQR) | 0.9 (0.7 to 1.0) | 1.7 (1.5 to 2.1) |
| Serum Cystatin-C (mg/L), median (IQR) | 0.9 (0.7 to 1.0) | 1.6 (1.4 to 2.0) |
| eGFR (mL/min/1.73 m<sup>2</sup>), CKD-EPI 2012 | 88.8 (17.1) | 37.6 (12.5) |
| eGFR (mL/min/1.73 m<sup>2</sup>), CKD-EPI 2021 | 91.1 (18.3) | 38.4 (12.3) |
| Urine albumin excretion rate (mg/24 hours), median (IQR) | 5.7 (3.5 to 8.5) | 39.2 (14.2 to 225.1) |
| CRP (mg/dL), median (IQR) | 0.1 (0.06 to 0.3) | 0.3 (0.1 to 0.7) |
| IL-6 (pg/mL), median (IQR) | 0.9 (0.6 to 1.4) | 1.5 (0.9 to 2.1) |
| TNF-α (pg/mL), median (IQR) | 1.6 (1.3 to 1.9) | 2.7 (2.1 to 3.0) |

Chronic kidney disease was defined as estimated glomerular filtration rate <60 ml/min per 1.73 m<sup>2</sup>; controls as ≥ 60 ml/min per 1.73 m<sup>2</sup>. Data are means (SDs) for continuous variables, N (percentages) for categorical variables, and medians (interquartile ranges). Abbreviations: SD, standard deviation; IQR, interquartile range; CVD, cardiovascular disease; HAP, human activity profile; eGFR, estimated glomerular filtration rate; CRP, C-reactive protein; IL-6, interleukin 6; TNF-α , tumor necrosis factor alpha

### Cross-sectional associations with total incretin levels (tAUC) and incretin response (iAUC) during OGTT in the overall cohort

The mean ± SD incremental and total GLP-1 area under the curve (GLP-1 iAUC and tAUC) during the OGTT were 1464 ± 1460 and 3043 ± 1899 pM x min respectively. The mean incremental and total post-prandial GIP area under the curve (GIP iAUC and tAUC) were 54327 ± 31785 and 68653 ± 36880 pg x min/ml respectively in the overall cohort (**Table 2)**. Both GLP-1 and GIP response (iAUC) were negatively correlated with caloric intake (r=-0.26 and -0.30; *P*<0.05), lean mass (r=-0.37 and -0.24, *P*<0.05) and physical activity (r=-0.17, *P*=0.08 and r=-0.28, *P*<0.05) in the overall cohort. In comparison we found no association of body composition or age with either total GLP-1 or GIP. In the overall cohort, eGFR was inversely correlated with only total GLP-1 levels (tAUC), but not GLP-1 response (iAUC) (**Figure 1A and 1C**). In the CKD subgroup, eGFR was inversely correlated with both total GLP-1 levels and GLP-1 response (r=-0.37 and r=-0.26 *P*<0.05). In comparison, eGFR was inversely correlated with both total GIP and GIP response in the overall cohort (**Figure 1B and 1D**). There was no significant or meaningful correlation of eGFR with total GIP (r=0.17, *P*=0.21) or iAUC (r=0.17, *P*=0.21) in the CKD subgroup.

**Figure 1.**
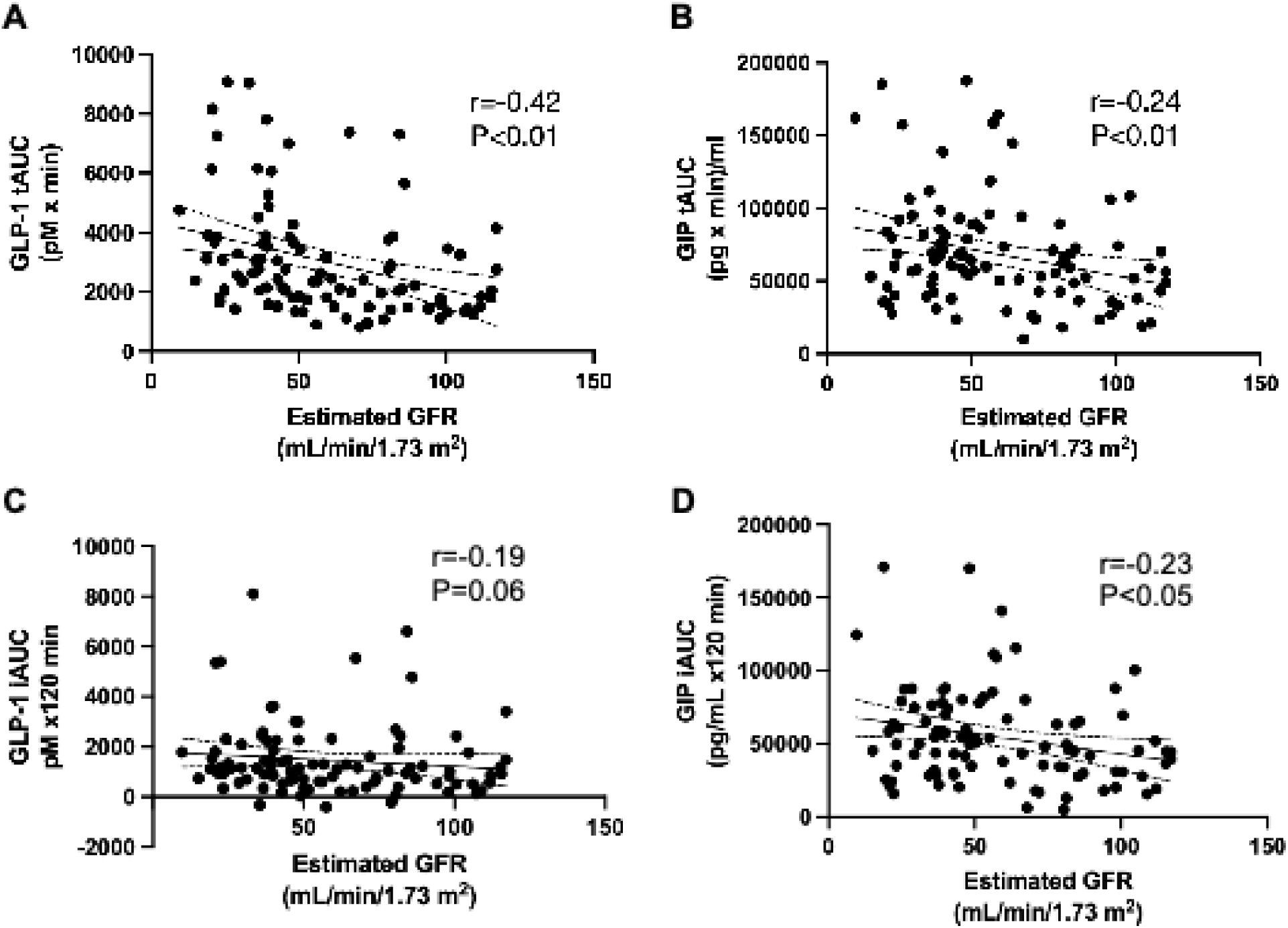
Association of estimated GFR with plasma incretin levels during OGTT. Distributions of GLP-1 and GIP response summarized using boxplots and scatterplots. eGFR<30 (n=17), eGFR 30-45 (n=22), eGFR 45-60 (n=19), and eGFR>60 (n=39). CKD-EPI creatinine-cystatin equation (2012) was used to estimate GFR. Spearman correlation coefficients were used to estimate the univariate relationship between incretin response and kidney function.

**Table 2.**
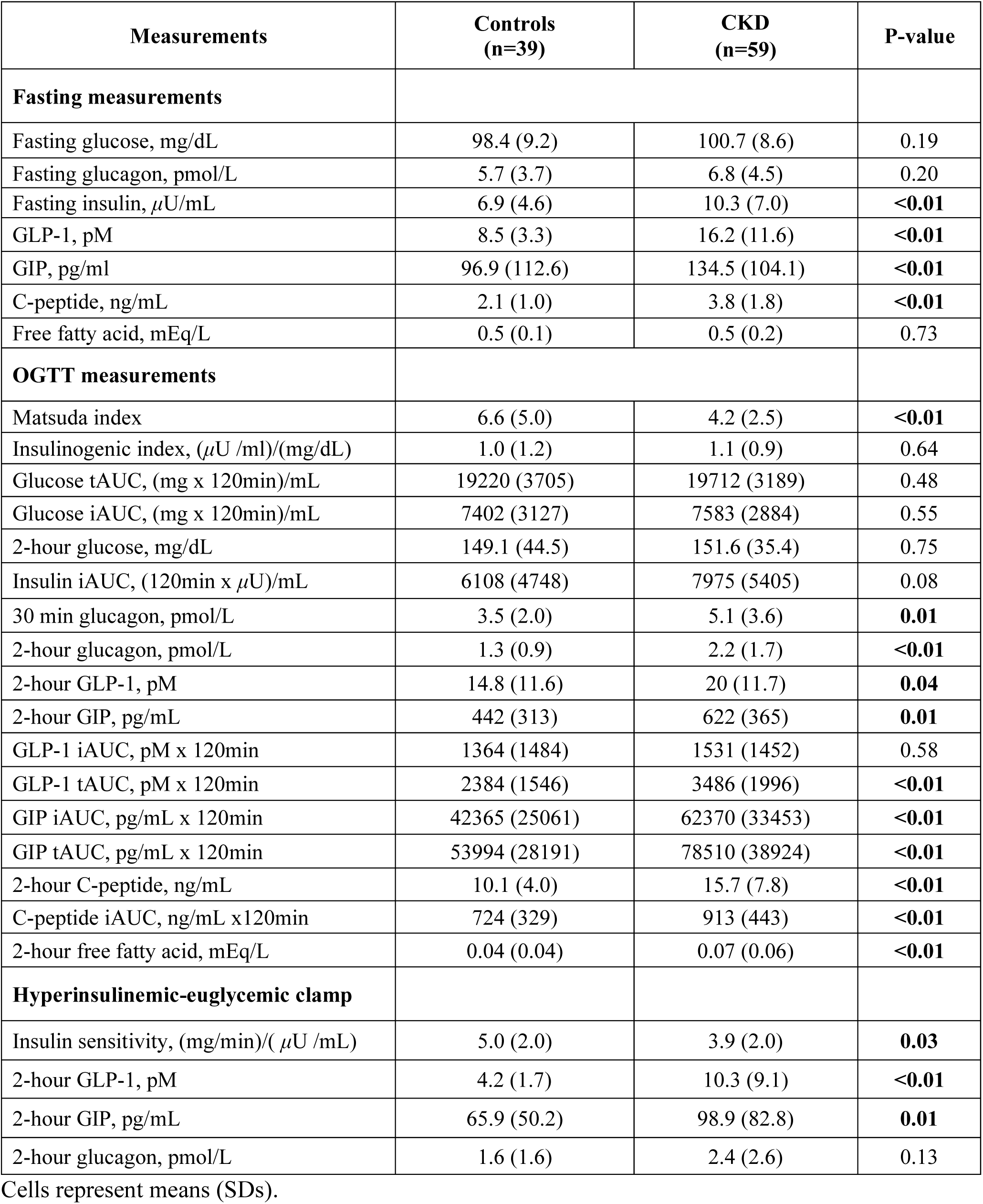
Fasting and OGTT glucose homeostasis and physiological measurements by CKD status.

| Measurements | Controls<br>(n=39) | CKD<br>(n=59) | P-value |
| --- | --- | --- | --- |
| **Fasting measurements** |  |  |  |
| Fasting glucose, mg/dL | 98.4 (9.2) | 100.7 (8.6) | 0.19 |
| Fasting glucagon, pmol/L | 5.7 (3.7) | 6.8 (4.5) | 0.20 |
| Fasting insulin, μU/mL | 6.9 (4.6) | 10.3 (7.0) | <0.01 |
| GLP-1, pM | 8.5 (3.3) | 16.2 (11.6) | <0.01 |
| GIP, pg/ml | 96.9 (112.6) | 134.5 (104.1) | <0.01 |
| C-peptide, ng/mL | 2.1 (1.0) | 3.8 (1.8) | <0.01 |
| Free fatty acid, mEq/L | 0.5 (0.1) | 0.5 (0.2) | 0.73 |
| **OGTT measurements** |  |  |  |
| Matsuda index | 6.6 (5.0) | 4.2 (2.5) | <0.01 |
| Insulinogenic index, (μU /ml)/(mg/dL) | 1.0 (1.2) | 1.1 (0.9) | 0.64 |
| Glucose tAUC, (mg x 120min)/mL | 19220 (3705) | 19712 (3189) | 0.48 |
| Glucose iAUC, (mg x 120min)/mL | 7402 (3127) | 7583 (2884) | 0.55 |
| 2-hour glucose, mg/dL | 149.1 (44.5) | 151.6 (35.4) | 0.75 |
| Insulin iAUC, (120min x μU)/mL | 6108 (4748) | 7975 (5405) | 0.08 |
| 30 min glucagon, pmol/L | 3.5 (2.0) | 5.1 (3.6) | 0.01 |
| 2-hour glucagon, pmol/L | 1.3 (0.9) | 2.2 (1.7) | <0.01 |
| 2-hour GLP-1, pM | 14.8 (11.6) | 20 (11.7) | 0.04 |
| 2-hour GIP, pg/mL | 442 (313) | 622 (365) | 0.01 |
| GLP-1 iAUC, pM x 120min | 1364 (1484) | 1531 (1452) | 0.58 |
| GLP-1 tAUC, pM x 120min | 2384 (1546) | 3486 (1996) | <0.01 |
| GIP iAUC, pg/mL x 120min | 42365 (25061) | 62370 (33453) | <0.01 |
| GIP tAUC, pg/mL x 120min | 53994 (28191) | 78510 (38924) | <0.01 |
| 2-hour C-peptide, ng/mL | 10.1 (4.0) | 15.7 (7.8) | <0.01 |
| C-peptide iAUC, ng/mL x120min | 724 (329) | 913 (443) | <0.01 |
| 2-hour free fatty acid, mEq/L | 0.04 (0.04) | 0.07 (0.06) | <0.01 |
| **Hyperinsulinemic-euglycemic clamp** |  |  |  |
| Insulin sensitivity, (mg/min)/(μU /mL) | 5.0 (2.0) | 3.9 (2.0) | 0.03 |
| 2-hour GLP-1, pM | 4.2 (1.7) | 10.3 (9.1) | <0.01 |
| 2-hour GIP, pg/mL | 65.9 (50.2) | 98.9 (82.8) | 0.01 |
| 2-hour glucagon, pmol/L | 1.6 (1.6) | 2.4 (2.6) | 0.13 |

Cells represent means (SDs).

### CKD was associated with greater fasting plasma incretin levels and varied incretin response during an OGTT

CKD was associated with a higher fasting GLP-1 levels with a mean of 16.2 ± 11.6 compared to 8.5 ± 3.3 pM among controls (*P*<0.01) (**Table 2, Supplemental Table 1**). GLP-1 tAUC measured during the OGTT was higher in participants with CKD versus controls (**Table 2**, **Figure 2A**). After adjusting for age, sex and race, CKD was associated with a 1192 pM x min higher GLP-1 tAUC (95% CI of 406 to 1978; *P*<0.01) (**Table 3**). Adjusting for other clinically relevant covariates only modestly attenuated the magnitude of the association (**Table 3**). In the final multivariable adjusted model, CKD was associated with a 1100 pM x min higher GLP-1 tAUC (95% CI of 119 to 2080; *P=*0.03) (**Table 3**). Despite CKD patients having higher total GLP-1 levels at fasting and during the OGTT, there was no significant difference in GLP-1 response (GLP-1 iAUC) compared to controls (**Tables 2 and Table 3**).

**Figure 2.**
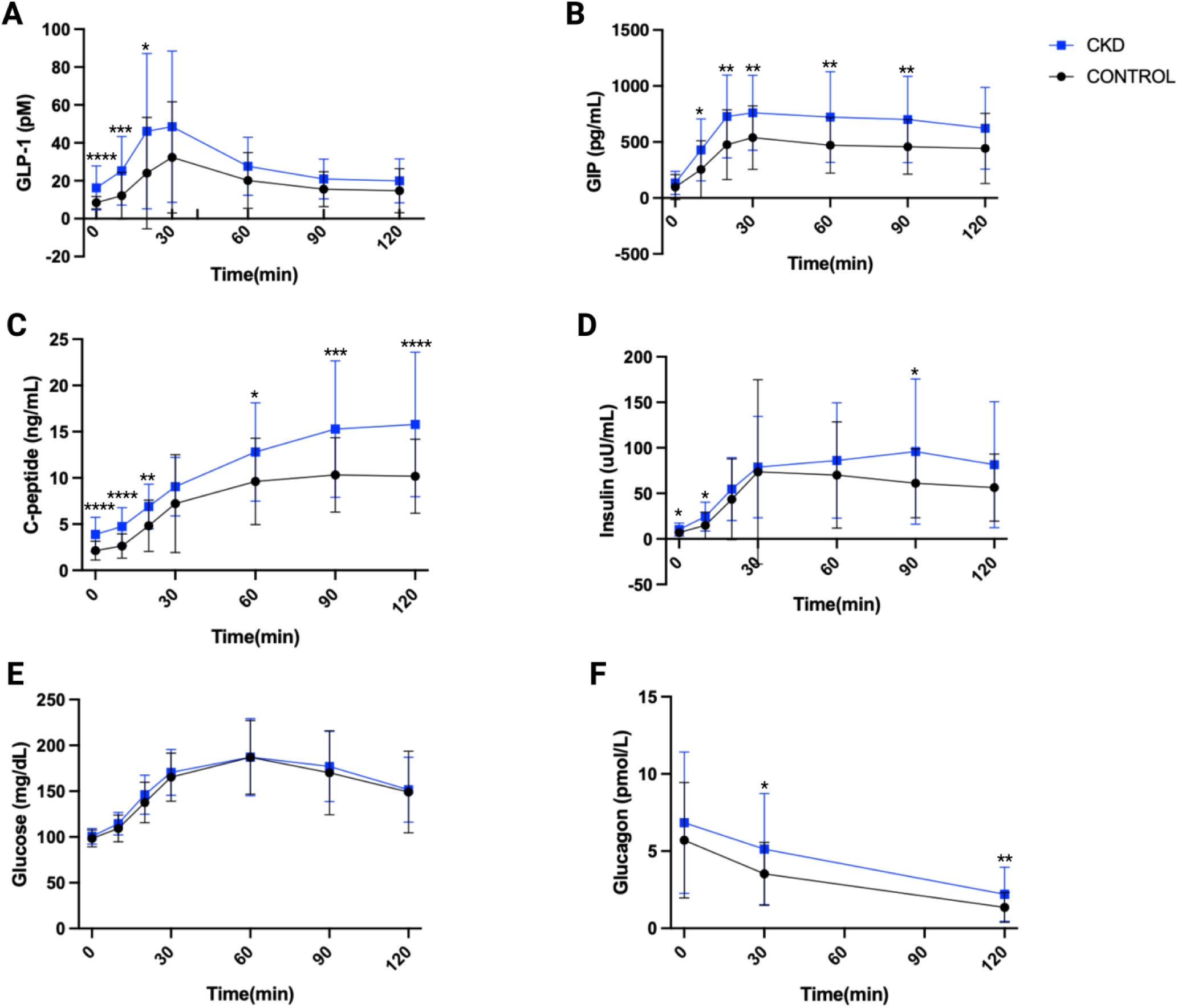
Changes in plasma glucose, glucagon, and pro-insulin factors in response to OGTT comparing CKD and controls. Data points and error bars are means and SD, respectively. One-way ANOVA corrected by multiple hypothesis testing (Bonferroni) was used to evaluate differences between CKD and controls at each timepoint. “****”= *P*<0.0001, “***” = *P*<0.001, “**” = *P*<0.01, “*” = *P*<0.05.

**Table 3.** Association of CKD with measures of GLP-1 and GIP during 2-hour OGTT. Mean differences represent the differences associated with CKD (vs controls) with 95% confidence intervals and P-values. Covariates were added one at a time to the base model which included age, sex, and race. The fully adjusted model is adjusted for age, sex, race, fat-free mass, fat mass, physical activity, calorie intake, smoking status and CVD. GLP-1 and GIP were measured during OGTT.

| Covariate adjustment | GLP-1 AUC |  |  |  | GIP AUC |  |  |  |
| --- | --- | --- | --- | --- | --- | --- | --- | --- |
|  | GLP-1 iAUC |  | GLP-1 tAUC |  | GIP iAUC |  | GIP tAUC |  |
|  | Difference (95% CI), pM x min | P | Difference (95% CI), pM x min | P | Difference (95% CI), (pg x min)/mL | P | Difference (95% CI), (pg x min)/mL | P |
| None (unadjusted) | 166 (-435 to 769) | 0.58 | 1102 (350 to 1854) | <0.01 | 20005 (7517 to 32493) | <0.01 | 24516 (10116 to 38916) | <0.01 |
| Age, sex, and race | 92 (-504 to 690) | 0.76 | 1192 (406 to 1978) | <0.01 | 18971 (5923 to 32018) | <0.01 | 21613 (6885 to 36340) | <0.01 |
| Weight | 162 (-447 to 771) | 0.60 | 1224 (417 to 2031) | <0.01 | 19629 (6244 to 33014) | <0.01 | 21908 (6783 to 37032) | <0.01 |
| Fat mass | 216 (-401 to 833) | 0.49 | 1223 (400 to 2045) | <0.01 | 19715 (5800 to 33630) | <0.01 | 21349 (5670 to 37029) | <0.01 |
| Fat-free mass | 144 (-515 to 803) | 0.66 | 1095 (217 to 1972) | 0.01 | 21408 (6540 to 36277) | <0.01 | 23465 (6720 to 40210) | <0.01 |
| Physical activity | 96 (-570 to 761) | 0.77 | 1019 (135 to 1903) | 0.02 | 19725 (4849 to 34600) | <0.01 | 21558 (4809 to 38308) | 0.01 |
| Calorie intake | 93 (-628 to 814) | 0.79 | 1022 (62 to 1981) | 0.04 | 14485 (-2.7 to 28972) | 0.05 | 15510 (-583 to 31603) | 0.06 |
| Smoking status | 92 (-634 to 818) | 0.80 | 1018 (53 to 1983) | 0.04 | 14490 (-100 to 29080) | 0.05 | 15528 (-677 to 31733) | 0.06 |
| Fully adjusted model | 122 (-619 to 864) | 0.74 | 1100 (119 to 2080) | 0.03 | 15271 (387 to 30154) | 0.04 | 16974 (515 to 33432) | 0.04 |

Mean fasting GIP level was higher among the CKD group with a mean of 134.5 ± 104.1 versus 97 ± 112.6 pg/ml in controls (*P*<0.01) (**Table 2, Supplemental Table 1**), but the estimated mean difference was not significant after adjusting for potential confounders (**Supplemental Table 1**). In contrast, both total postprandial GIP level and GIP response were elevated in CKD compared to controls (**Table 2 and Figure 2B**). Adjusting for potential confounders attenuated the estimated association by 24% to an estimated mean difference of 15271 pg x min/ml higher GIP iAUC (95% CI of 387 to 30154; *P=*0.04) in CKD compared to controls (**Table 3**).

The rate of acute GIP increase in the first 30 minutes of OGTT was greater in CKD compared to controls. The mean rate of increase in GIP within the first 30 minutes of the OGTT was 249 ± 111 vs 177 ± 101 pg/ml/min in CKD and controls, respectively. CKD patients had an estimated mean 167pg/ml/min greater rate of increase in GIP (95% CI of 50 to 284; P<0.01) compared to controls after adjustment for potential confounders (**Supplemental Table 2**). Further adjustment for fasting plasma GIP levels did not meaningfully impact estimates of association. In contrast, the CKD patients did not differ meaningfully or significantly in their mean rate of increase in GLP-1 during the first 30 minutes of the OGTT (**Supplemental Table 2**).

### GIP response, but not GLP-1 response was associated with insulinotropic effects during OGTT

Total postprandial insulin levels during the OGTT did not significantly differ between CKD and controls, whereas C-peptide levels were more consistently greater at each time point in CKD during the OGTT **(Figure 2C and 2D)**. No significant differences were observed in insulin response measured by insulin iAUC and insulinogenic index between CKD and controls (Table 2). Similarly, we found no meaningful or significant difference by CKD status in glucose tolerance measured by glucose iAUC (**Table 2**, **Figure 2E).** GLP-1 response (GLP-1 iAUC) was not meaningfully or significantly associated with insulin, C-peptide, or glucose iAUCs in the overall cohort (**Supplemental Figure 1A, 1C, and 1E**). In the overall cohort, GIP response (GIP iAUC) was significantly correlated with insulin (r=0.25, *P* =0.01) and C-peptide response (0.29, *P* <0.01) but not glucose iAUC (r=-0.03, *P*=0.78). These correlations were generally weaker in patients with CKD (r=0.21, *P*=0.12; r=0.24, *P*=0.07; r=0.03, *P*=0.92, respectively) compared with controls (r=0.33, *P*=0.04; r=0.47, *P*<0.01; r=-0.17, *P*=0.29, respectively) (**Supplemental Figure 1B, 1D and 1F**).

### Plasma glucagon levels were elevated in CKD compared to controls in response to OGTT

Fasting plasma glucagon levels were not significantly different between CKD and controls (**Table 2, Supplemental Table 1**, **Figure 2F)**. During the OGTT plasma glucagon levels were higher at 30 minutes and 120 minutes in CKD compared to controls (**Table 2**, **Figure 2F**). After adjusting for baseline glucagon levels, CKD was associated with 0.9 mg/dl higher levels at 30 minutes (95% CI of 0.15, 1.7; *P*=0.02) and 0.5 mg/dl higher at 120 minutes (95% CI of 0.1 to 0.9; *P*=0.02) post OGTT. The percent change in glucagon levels from baseline to 30 minutes post OGTT was attenuated in CKD with a median [IQR] of -27% [-11 to -46] versus -38% [-19 to -57] among controls. The percent change from baseline was also modestly attenuated at 2 hours post OGTT among CKD with median [IQR] of -70% [-57 to -80] compared to -78% [-60 to -88] in controls.

### The fasting plasma dipeptidyl peptidase-4 (DPP-4) activity and antigen levels were similar between CKD and controls

The mean fasting plasma DPP-4 antigen levels were similar among CKD and controls (mean ± SD= 56.2 ± 15.3 versus 55.7 ± 15.5 ng/ml; *P*=0.88) (**Figure 3A**). The mean fasting plasma DPP-4 activity levels were also similar among the two groups (mean ± SD= 28.7 ± 7.3 versus 28.8 ± 6.3 µM/min; *P*=0.95) (**Figure 3B**).

**Figure 3.**
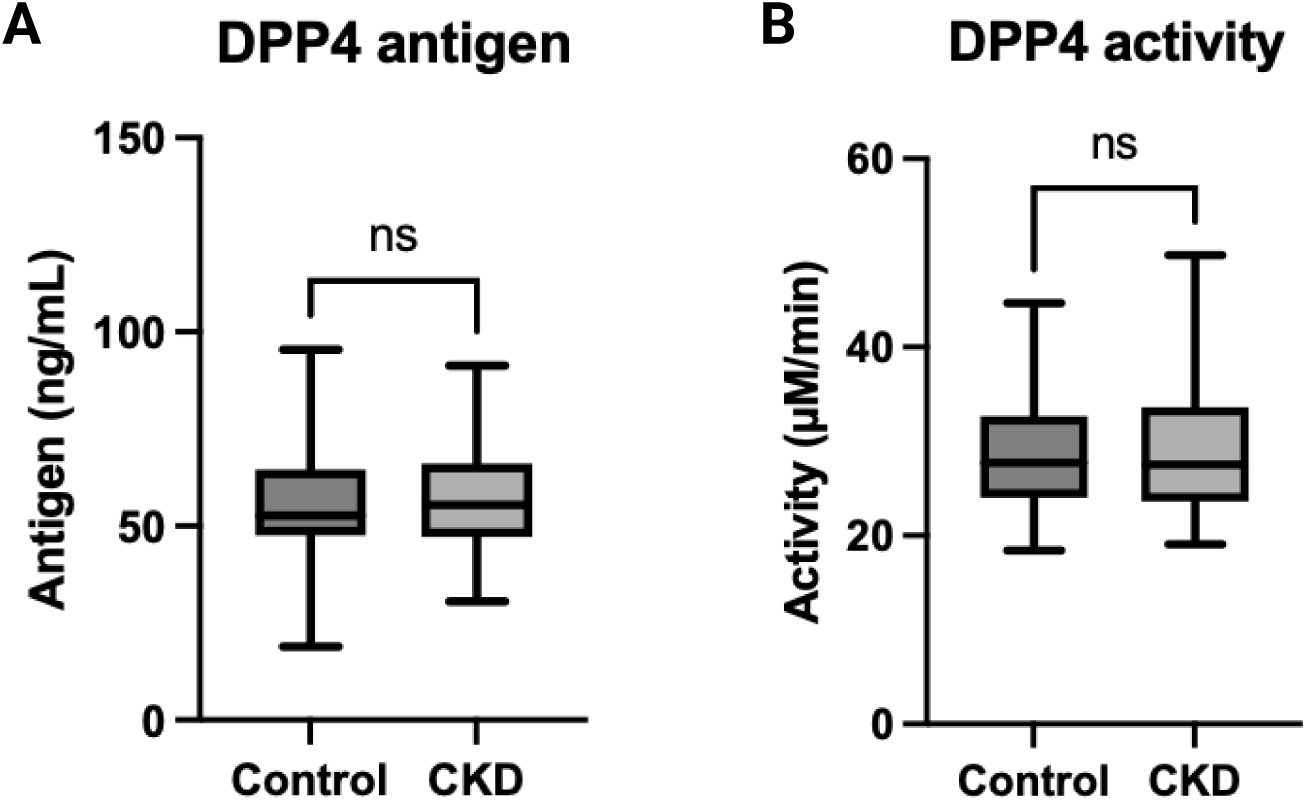
Comparison of fasting plasma DPP-4 antigen and activity levels among CKD (n=43) and controls (n=34). Box plots represent median and IQR and the whiskers represent minimum and maximum values. Unpaired t-Test was used to determine the difference between the two groups.

### Greater inflammation was associated with greater incretin levels and incretin response in CKD

In the overall cohort, plasma TNF-α levels were significantly associated with GIP response, and CRP levels were significantly associated with GLP-1 response (**Supplemental Table 3**). In the CKD subgroup, greater CRP was also associated with greater GLP-1 response (**Supplemental Table 3**). Among patients with CKD each 1 mg/dL greater plasma CRP was associated with 0.58 greater pM GLP-1 response (95% CI of 0.37 to 0.8; *P*<0.01) in CKD (**Supplemental Table 3**).

### Sensitivity analyses using the CKD-EPI creatinine-cystatin C 2021 equation yielded similar outcomes

Using the race-neutral equation, three CKD participants were reclassified to controls resulting in 42 controls and 56 CKD. The eGFR was similar among CKD and controls compared to the 2012 equation (**Table 1**). Despite the modest shift in group assignments, all the above analyses were replicated with the 2021 formula and showed similar outcomes (**Supplemental Table 4 and Supplemental Figure 2**).

## Discussion

Our findings demonstrate that the presence and severity of non-diabetic moderate-severe CKD is associated with greater plasma levels of incretins during fasting and in response to an OGTT. The elevated circulating GLP-1 and GIP levels in the fasting state and postprandial conditions were observed in the absence of any significant difference in fasting glucagon levels, DPP-4 antigen, or activity levels. Acute GIP release and GIP response (iAUC) during the OGTT were significantly higher in CKD compared to controls. The correlation of incretin levels with OGTT stimulated insulin or c-peptide was attenuated in those with CKD compared with controls. Concomitantly, CKD was associated with elevated postprandial plasma glucagon levels and impaired glucagon suppression post OGTT. In CKD, the inflammatory biomarker CRP was associated with elevated incretin response. Overall, our findings show that non-diabetic moderate-severe CKD is associated with greater postprandial incretin levels and an augmented GIP response during OGTT that do not translate into meaningful improvements in insulin, glucose, or glucagon homeostasis.

We found elevated fasting and post-prandial plasma incretin levels in CKD was independent of differences in circulating fasting DPP-4 levels and activity suggesting that these differences are unlikely due to reduced incretin degradation. DPP-4 is considered the predominant enzyme responsible for incretin degradation, however it remains unknown if DPP-4 activity is altered during oral glucose tolerance testing in CKD. It is notable that our findings are consistent with other studies in patients with non-diabetic end-stage renal disease (ESRD). One prior study showed greater GLP-1 levels in response to a high-calorie mixed meal in non-diabetic end-stage renal disease (ESRD) subjects compared to healthy controls^20^ while another small study of nine non-diabetic hemodialysis patients and 10 healthy controls found elevated fasting and postprandial total GIP response during a standardized meal^21^. Like these prior studies, we measured only total GLP-1 and GIP and are unable to distinguish the proportion of active from inactive incretin fragments in our CKD patients. Future studies are needed to confirm if the augmented incretin levels reflect parallel increases active GLP-1 and GIP secretion in CKD and the possible influence of the uremic milieu on potential alternative incretin degradation pathways.

In our study, CKD was associated with a greater rate of GIP increase but not GLP-1 increase in the first 30 minutes of OGTT compared to controls (**Supplemental Table 2**). This difference in the rate of GIP increase between CKD and controls was independent of differences in fasting levels of GIP implying that these differences may be independent of reduced clearance of GIP. Controversy exists regarding the role of renal clearance on incretin response. A prior small case-control study in a select group of patients with more modest kidney disease (mean creatinine clearance 46 ml/min) suggested similar metabolic clearance rates and plasma half-life of intact GLP-1 and intact GIP but prolonged metabolite half-lives with intravenous GLP-1 and GIP infusion in CKD compared to controls^22^. This study was limited by both the lack of any urinary measurements necessary to accurately assess renal clearance and lack of assessment of lean mass known to be reduced in patients with CKD influencing the volume of distribution and confounding estimates of drug clearance. Another study in patients with ESRD treated with dialysis showed no difference in incretin response compared with controls casting doubt on the impact of renal clearance on incretin response suggesting a preserved ability to degrade and eliminate active GLP-1 and GIP and their metabolites in ESRD ^23^. More detailed studies are needed to directly assess secretion, elimination, and breakdown of intact incretin hormones and their metabolites across the spectrum of CKD.

Disruption of postprandial incretin hormone response (iAUC) in CKD appeared to influence downstream insulin, c-peptide and glucagon homeostasis during the OGTT. In healthy adults, GIP is considered more strongly insulinotropic than GLP-1^24^. Consistent with these findings we found a stronger positive correlation between GIP response and insulin/C-peptide compared to GLP-1. Furthermore, we noted that this correlation between GIP response and insulin/C-peptide was noticeably weaker in patients with CKD compared to controls. In comparison, we found no meaningful correlation of GLP-1 with insulinotropic response. Our findings expand on those of prior studies suggesting that non-diabetic patients with CKD demonstrate a blunted insulinotropic effect of incretins akin to patients with type 2 diabetes and normal kidney function ^25,26^. However, CKD patients appeared to have numerically greater baseline-corrected insulin response (insulin iAUC) reflecting reduced insulin clearance^3^ and a similar acute insulin response estimated by the insulinogenic index compared to controls (**Table 2**). This may suggest that altered glucose homeostasis in CKD patients may be attributed to inadequate augmentation of the insulin response by incretin hormones (especially GLP-1) or resistance to insulin’s actions on peripheral tissues. Our findings are consistent with results from a randomized double-blind study that also showed that non-diabetic ESRD patients exhibit reduced incretin action on insulin production of both GLP-1 and GIP despite adequate insulin response during IV glucose stimulation^27^. While mechanistic studies of CKD in 5/6^th^ nephrectomized mice have observed impaired β-cell insulin secretion in response to glucose ^28^, none have specifically investigated β-cell resistance to GIP activity on insulin secretion. These findings motivate mechanistic studies to investigate if disruption in the incretin response to carbohydrate consumption in non-diabetic CKD reflects resistance to incretin hormones, especially in the β-cells of the endocrine pancreas where GLP-1 and GIP receptors are abundantly expressed ^29^.

The attenuated suppression of glucagon during the OGTT in non-diabetic moderate-severe CKD observed in our study also suggests potential disruption of alpha cell response to incretins in CKD. Despite declines in glucagon levels during the OGTT in both CKD and controls, postprandial glucagon levels remained significantly higher in the CKD group compared to controls. These findings are in line with other studies of patients with type 2 diabetes and non-diabetic patients with ESRD^5,23,30–32^. It suggests an altered counterregulatory balance between GIP induction and GLP-1 suppression of alpha cell glucagon production in CKD during OGTT-induced hyperglycemia. Sustained and elevated postprandial glucagon levels could have direct adverse impacts on glycemic control and amino acid catabolism contributing to muscle wasting in patients with CKD^33–35^. Further studies are needed to assess the factors contributing to postprandial glucagon hypersecretion and inadequate suppression and its contribution to metabolic dysregulation in CKD.

Inflammation was identified as a contributing factor for heightened incretin response to OGTT. We found that plasma C-reactive protein (CRP) (a marker for systemic inflammatory burden) was significantly associated with GLP-1 response during OGTT in CKD independent of other factors (**Table 4**). The association of inflammatory biomarkers including CRP and IL-6 with GLP-1 levels has been reported in other observational studies ^36–38^. Evidence from studies of patients in the intensive care unit show a significant association of greater inflammatory biomarkers levels including IL-6 and CRP and GLP-1^36^. This suggests a crosstalk between inflammatory status associated with CKD and glucose metabolism regulation through gut-driven incretin response. Interestingly the contrary has been observed with administration of exogenous incretin mimetic therapies associated with a strong anti-inflammatory response. Studies have shown long-term incretin-based therapies significantly decreases in circulating proinflammatory cytokines, including IL-6, TNF-α, IL-1*β*, and MCP-1 ^39–41^. Future studies are needed to determine the biological mechanism linking elevated endogenous incretin levels and systemic inflammation in CKD and if treatment with incretin analogues may influence inflammation and catabolism in CKD.

Our study had notable strengths and limitations. First, we recruited a relatively large group of well-characterized non-diabetic CKD participants across the spectrum of moderate-severe CKD including measures of body composition and lifestyle factors. Second, we used an OGTT to comprehensively measure gut-derived incretin hormones, glucagon, insulin, and glucose. Third, we employed a rigorous analysis method adjusting for a wide range of potential confounders in the association of CKD with circulating incretin levels and incretin response to oral glucose. Our study was not without limitations. First, our assays measured total GLP-1 and GIP levels in the plasma, so the proportion of active from the total GLP-1 and GIP and their renal clearance was not directly measured. Second, despite normal fasting glucose levels, both controls and CKD patients included individuals with impaired glucose tolerance (IGT) defined by 2-hour glucose level 140mg/dL or above. However, the inclusion of individuals with IGT in our control group may suggest that the estimated differences in incretin levels and response are conservative. Third, serial blood sample collections during OGTT and clamp were acquired without the addition of a DPP-4 inhibitor which may have impacted the levels of detected glucagon, GLP-1 and GIP. We addressed this by measuring both the plasma fasting DPP-4 antigen levels and its activity and found similar antigen and activity levels among both groups.

In conclusion, non-diabetic CKD is associated with disruption of incretin homeostasis and evidence of attenuated incretin effects on insulin, C-peptide, and glucagon secretion. These changes may contribute to the metabolic dysregulation associated with kidney disease and reveal a potential role for incretin-mimetics to address the attenuated incretin effects observed in our study. Indeed, a recent pharmacokinetic study of combination GLP-1 and GIP in the form of single-dose tirzepatide, a dual GLP-1 and GIP receptor agonist, showed similar drug clearance and tolerability in healthy controls compared to patients across all stages of CKD, including ESRD ^42^. Studies are needed to investigate the differential efficacy of GLP-1 and GIP single and dual agonist on insulin, glucose and glucagon homeostasis and links to outcomes in non-diabetic CKD.

## Data Availability

All data produced in the present study are available upon reasonable request to the authors

## Disclosures

The authors have no relevant disclosures to report.

## Funding

Funding for this study was provided by an unrestricted grant from the Northwest Kidney Centers and R01DK087726 (IHDB), R01DK087726-S1 (IHDB), R01DK129793 (BR), R01DK087726, R01DK087726-S1, K01 DK102851 (JAA, IHDB), R01DK125794 (JLG), K24 DK096574 (TRZ), R56DK124853 (BPC), and P30 DK017047 (University of Washington Diabetes Research Center) and Dialysis Clinics Incorporated, C-4122 (BR).

## Author Contributions

The conceptualization was contributed by AA, BR, BPC, and IHDB. The methodology was contributed by AA, BR, BPC, BC, SF and IHDB. The formal analysis was conducted by AA and SF. The investigation was performed by BR, JG, BPC, and IHDB. Resources were contributed by BR, BPC, JG, and IHDB. Data curation was performed by AA, SF, MT, and LRZ. The original draft was written by AA and BR. The review was written and edited by AA, BR, BL, BJB, JG, JEN, BE, IHDB, BJB, JH, BPC, MT and LFB. Visualization was contributed by AA and SF. Supervision was carried out by BR, BRK, and BPC. Project administration was contributed by BR, JG, IHDB, and BPC. Funding acquisition was contributed by BR, JG, BPC, and IHDB.

## Data sharing statement

Deidentified data, which have been stripped of all personal identification and information, will be made available to share upon request as part of the research collaboration.

## Acknowledgments

We thank Anthony Dematteo at Vanderbilt University who measured plasma DPP-4 antigen and activity levels. We would like to express our sincere gratitude to Steven Kahn for his valuable feedback and insightful comments on this manuscript. We thank all the participants in the SUGAR cohort for their contributions to this investigation.

---

**Supplemental Figure 1.**
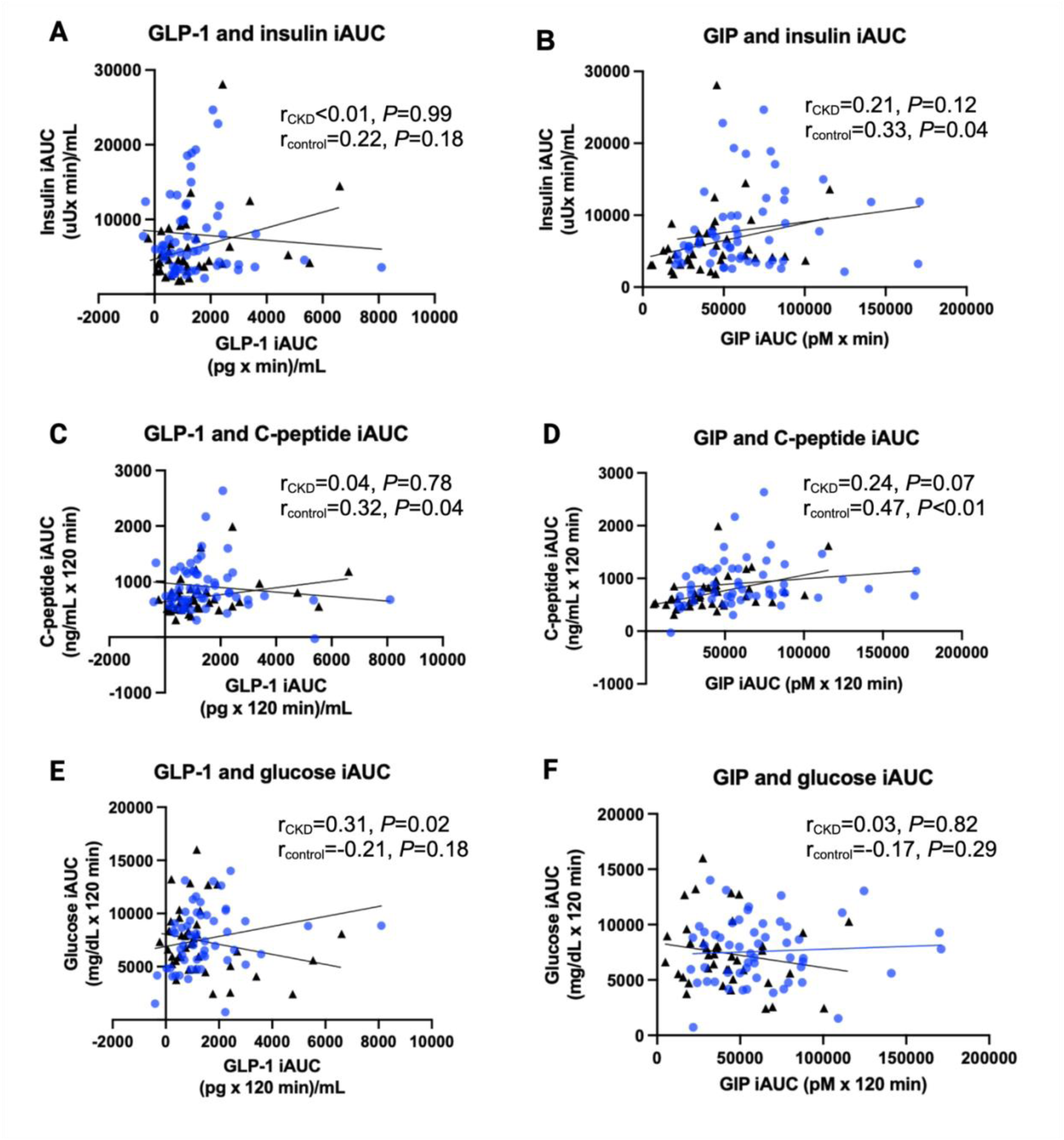
The correlation between incretin response with insulin, C-peptide, and, glucose iAUCs in CKD and controls during OGTT. Spearman correlation coefficient was used to estimate the univariate relationship between incretin response and insulin secretion. Black triangles represent controls and blue circle represent CKD.

**Supplemental Figure 2.**
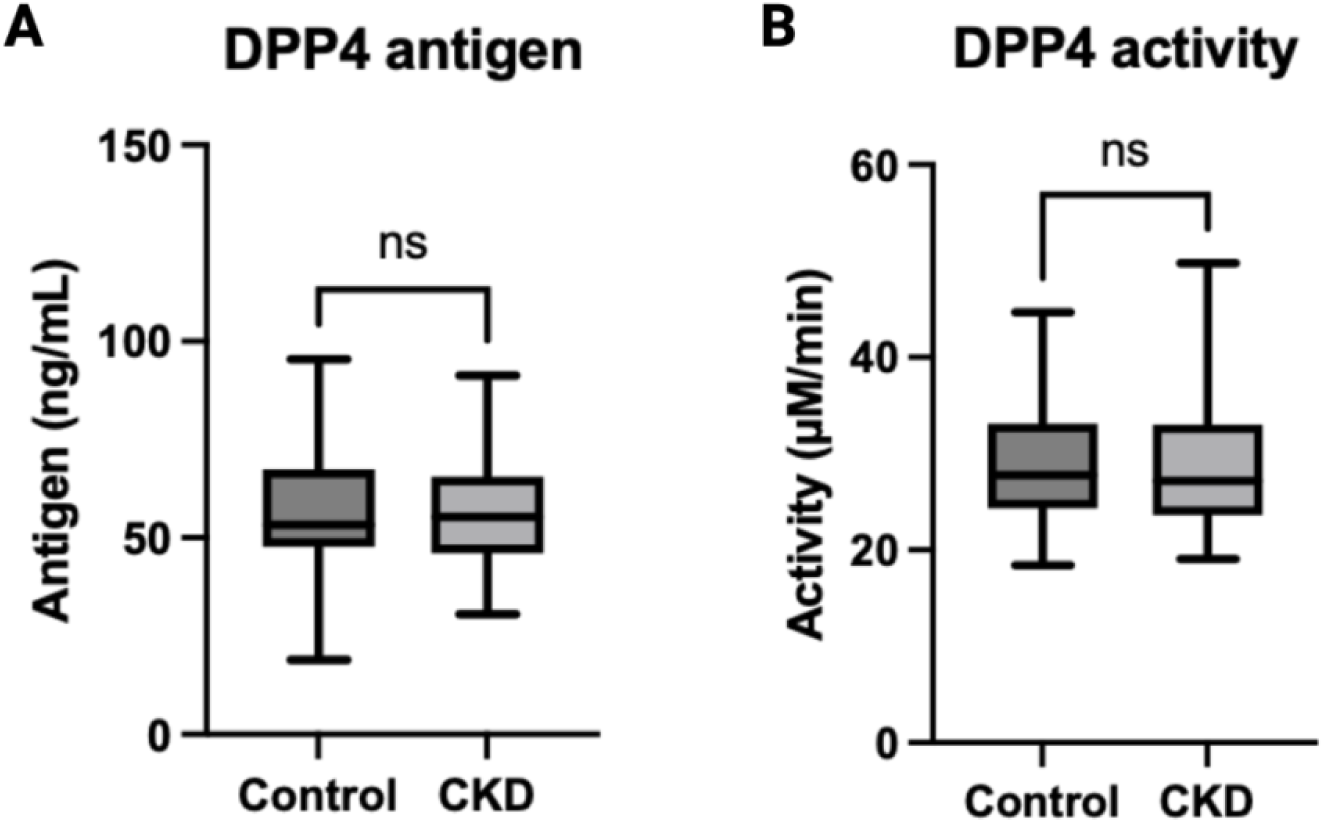
Comparison of fasting plasma DPP4 antigen and activity levels among CKD (n=41) and controls (n=36) using the CKD-EPI Creatinine-Cystatin C Equation (2021). Box plots represent median and IQR and the whiskers represent minimum and maximum values. Unpaired t-Test was used to determine the difference between the two groups.

**Supplemental Table 1.** Association of CKD with fasting GLP-1, GIP, and glucagon measurements. Mexan differences represent the differences associated with CKD (vs controls) with 95% confidence intervals and P-values. Covariates were added one at a time to the base model which included age, sex, and race. The fully adjusted model is adjusted for age, sex, race, fat-free mass, fat mass, physical activity, calorie intake, smoking status, and CVD.

| Covariate adjustment | Fasting GLP-1 |  | Fasting GIP |  | Fasting glucagon |  |
| --- | --- | --- | --- | --- | --- | --- |
|  | Difference (95% CI), pM | *P* | Difference (95% CI), pg/mL | *P* | Difference (95% CI), pmol/L | *P* |
| None (unadjusted) | 8.8 (5.1 to 12) | <0.01 | 52 (18 to 85) | <0.01 | 1.1 (-0.6 to 2.9) | 0.20 |
| Age, sex, and race | 10.1 (6.2 to 14) | <0.01 | 36 (4.3 to 68) | 0.03 | 1.7 (0.03 to 3.4) | 0.04 |
| Weight | 9.9 (5.8 to 14) | <0.01 | 35 (2.2 to 67) | 0.04 | 1.6 (-0.1 to 3.3) | 0.06 |
| Fat mass | 9.4 (5.2 to 13) | <0.01 | 31 (-2.6 to 65) | 0.07 | 1.5 (-0.2 to 3.3) | 0.09 |
| Fat-free mass | 8.7 (4.3 to 13) | <0.01 | 30 (-6.5 to 66) | 0.11 | 1.3 (-0.6 to 3.2) | 0.17 |
| Physical activity | 8.6 (4.1 to 13) | <0.01 | 29.5 (-7.3 to 66) | 0.11 | 1.1 (0.8 to 3.1) | 0.24 |
| Calorie intake | 8.5 (3.7 to 13) | <0.01 | 24 (-13 to 61) | 0.20 | 0.9 (-1.1 to 2.9) | 0.36 |
| Smoking status | 8.5 (3.7 to 13) | <0.01 | 25.2 (-12 to 62) | 0.18 | 0.8 (-1.1 to 2.8) | 0.40 |
| Fully adjusted model | 8.8 (3.9 to 14) | <0.01 | 30 (-7.7 to 67) | 0.11 | 1.0 (-0.9 to 3.0) | 0.30 |

**Supplemental Table 2.** Estimated differences in the rate of acute incretin peripheral response between CKD and controls. Mean differences represent the differences associated with CKD (vs controls) with 95% confidence intervals and *P*-values. Covariates were added one at a time to the base model which included age, sex, weight, and smoking status.

| Covariate adjustment | Acute GLP-1 response |  | Acute GIP response |  |
| --- | --- | --- | --- | --- |
|  | Difference (95% CI), pM/min | *P* | Difference (95% CI), pg/mL/min | *P* |
| None (unadjusted) | 7.5 (-6.4 to 21.3) | 0.29 | 174 (61 to 287) | <0.01 |
| Age, sex, weight, smoking status | 7.5 (-6.4 to 21.3) | 0.29 | 172 (58 to 285) | <0.01 |
| Fat mass, fat-free mass, calorie intake | 10.6 (-4.1 to 25.3) | 0.16 | 167 (50 to 284) | <0.01 |
| Physical activity | 10.7 (-4.0 to 25.3) | 0.16 | 167 (50 to 284) | <0.01 |

**Supplemental Table 3.** Association of inflammatory biomarkers with incretin response during OGTT in the CKD and controls. Cells represent association coefficients with 95% confidence intervals and *P*-values. The coefficients represent the estimated change in incretin response with one unit increase in the corresponding inflammatory biomarker. Association coefficients were obtained using linear regression adjusting for adjusted for age, sex, race, fat-mass, fat-free mass, calorie intake, physical activity, and smoking status. * Significant association of inflammatory biomarker with GLP-1 or GIP response in the overall cohort. “**” = *P*<0.01, “*” = *P*<0.05.

| Inflammatory biomarker | GLP-1 iAUC |  |  |  | GIP iAUC |  |  |  |
| --- | --- | --- | --- | --- | --- | --- | --- | --- |
|  | Overall | Control | CKD | *P* for interaction | Overall | Control | CKD | *P* for interaction |
|  | β (95% CI) | β (95% CI) | β (95% CI) |  | β (95% CI) | β (95% CI) | β (95% CI) |  |
| C-reactive protein ** (mg/dL) | 0.29 (-0.1 to 0.5) | 0.04 (-0.26 to 0.35) | 0.58 (0.38 to 0.78)** | <0.01 | -0.06 (-4.4 to 4.3) | 0.83 (-4.7 to 6.4) | 0.5 (-6.3 to 7.2) | 0.88 |
| IL-6 (pg/mL) | -0.08 (-0.4 to 0.2) | -0.08 (-0.40 to 0.23) | -86 (-336 to 165) | 0.32 | 5.7 (-0.1 to 11.5) | 6.1 (1.0 to 11) | 2155 (-4074 to 8384) | 0.28 |
| IFN- γ (pg/mL) | 1.0 (-6.1 to 8.3) | 170 (-61 to 401) | 4.1 (-2.8 to 11) | 0.42 | 36 (-109 to 181) | 1770 (-2514 to 6054) | 34 (-141 to 209) | 0.29 |
| IL-1β (pg/mL) | -0.03 (-0.33 to 0.27) | -0.17 (-0.5 to 0.16) | 410 (-4594 to 5415) | 0.73 | -2.8 (-9.0 to 3.3) | -1.5 (-7.4 to 4.4) | 43070 (-80512 to 166653) | 0.34 |
| TNF-α* (pg/mL) | 186 (-204 to 575) | 283 (-651 to 1216) | 441 (-136 to 1020) | 0.88 | 8139 (370 to 15907)* | -4659 (-21511 to 12191) | 12223 (-2042 to 26489) | 0.11 |

**Supplemental Table 4.** Association of CKD with measures of GLP-1 and GIP response during 2-hour OGTT using the CKD-EPI Creatinine-Cystatin C Equation (2021). Mean differences represent the differences associated with CKD (vs controls) with 95% confidence intervals and P-values. Covariates were added one at a time to the base model which included age, sex, and race. The fully adjusted model is adjusted for age, sex, race, fat-free mass, fat mass, physical activity, calorie intake, smoking status, and CVD.

| Covariate adjustment | GLP-1 exposure |  |  |  | GIP exposure |  |  |  |
| --- | --- | --- | --- | --- | --- | --- | --- | --- |
|  | GLP-1 iAUC |  | GLP-1 tAUC |  | GIP iAUC |  | GIP tAUC |  |
|  | Difference (95% CI), pM x min | P | Difference (95% CI), pM x min | P | Difference (95% CI), (pg x min)/mL | P | Difference (95% CI), (pg x min)/mL | P |
| None (unadjusted) | 233 (-362 to 828) | 0.44 | 1187 (449 to 1925) | <0.01 | 17900 (5424 to 30377) | <0.01 | 23054 (8722 to 37387) | <0.01 |
| Age, sex, and race | 224 (-364 to 814) | 0.45 | 1326 (559 to 2094) | <0.01 | 17244 (4234 to 30254) | <0.01 | 20281 (5633 to 34930) | <0.01 |
| Weight | 268 (-325 to 861) | 0.37 | 1336 (558 to 2115) | <0.01 | 17432 (4244 to 30621) | 0.01 | 20232 (5379 to 35086) | <0.01 |
| Fat mass | 325 (-274 to 925) | 0.28 | 1341 (549 to 2132) | <0.01 | 17452 (3760 to 31144) | 0.01 | 19676 (4303 to 35048) | 0.01 |
| Fat-free mass | 272 (-366 to 910) | 0.40 | 1234 (393 to 2075) | <0.01 | 18719 (4144 to 33294) | 0.01 | 21406 (5054 to 37757) | 0.01 |
| Physical activity | 226 (-419 to 871) | 0.49 | 1163 (314 to 2011) | <0.01 | 17007 (2422 to 31592) | 0.02 | 19487 (3124 to 35849) | 0.02 |
| Calorie intake | 235 (-462 to 932) | 0.50 | 1077 (259 to 2095) | 0.01 | 12043 (-2090 to 26177) | 0.09 | 13949 (-1673 to 29613) | 0.08 |
| Smoking status | 235 (-466 to 937) | 0.50 | 1178 (256 to 2101) | 0.01 | 12043 (-2188 to 26275) | 0.09 | 13966 (-1784 to 28718) | 0.08 |
| Fully adjusted model | 284 (-439 to 1007) | 0.43 | 1304 (360 to 2248) | <0.01 | 13099 (-1566 to 27764) | 0.08 | 15931 (-217 to 32079) | 0.05 |

## Notes

### Competing Interest Statement

The authors have declared no competing interest.

### Author Declarations

The procedures in the study and informed consent forms were reviewed and approved by the University of Washington Human Subjects Division (HSD).

